# In major depression, complex intersections between adverse childhood experiences and negative life events impact neuroticism, brooding, suicidal tendencies, insomnia, and cognitive interference

**DOI:** 10.1101/2024.01.04.24300880

**Authors:** Asara Vasupanrajit, Michael Maes, Ketsupar Jirakran, Chavit Tunvirachaisakul

**Author notes:** **Corresponding author:** Prof. Dr. Michael Maes, M.D., PhD. Department of Psychiatry, Faculty of Medicine, Chulalongkorn University, Bangkok, 10330, Thailand And Prof. Dr. Michael Maes, M.D., Ph.D. Sichuan Provincial Center for Mental Health Sichuan Provincial People’s Hospital, School of Medicine, University of Electronic Science and Technology of China Chengdu, 610072, China Michael Maes. Joint first authorship (Asara Vasupanrajit, Michael Maes). **Co-corresponding author:** Dr. Chavit Tunvirachaisakul, M.D., PhD. Department of Psychiatry, Faculty of Medicine, Chulalongkorn University, Bangkok, 10330, Thailand. **E-mail addresses:** Asara Vasupanrajit Ketsupar Jirakran Chavit Tunvirachaisakul Michael Maes.

## Abstract

**Background:** There is evidence that adverse childhood experiences (ACEs) and negative life events (NLEs) are associated with major depression (MDD).

**Purpose:** To determine whether ACEs affect all features of mild MDD, including suicidal tendencies, brooding, neuroticism, insomnia, cognitive deficits, severity of depression and anxiety, and cognitive deficits, and whether NLEs mediate these effects.

**Patients and methods:** This study examines a cohort of 118 academic students, namely 74 students who satisfied the DSM-5-TR criteria for MDD and 44 normal control students. We assessed brooding, neuroticism, suicidal ideation and attempts, and the severity of depression, anxiety, insomnia, and the Stroop tests.

**Results:** One validated factor could be extracted from brooding, neuroticism, current suicidal behaviors, and the severity of depression, anxiety, and insomnia, labeled the phenome of depression. A large part of the variance in the phenome of depression (55.0%) was explained by the combined effects of self-, relationships, and academic-related NLEs in conjunction with ACEs, including family dysfunction and abuse and neglect (both physical and emotional). The latter ACEs significantly interacted (moderating effect) with NLEs to impact the depression phenome. Although sexual abuse did not have direct effects on the phenome, its effects were mediated by NLEs. We discovered that increased sexual abuse, physical and emotional abuse and neglect, and ACEs related to family dysfunction predicted 22.5% of the variance in NLEs. Up to 18.5% of the variance in the Stroop test scores was explained by sexual abuse and the phenome of depression. The latter mediated the effects of NLEs and abuse, neglect, and family dysfunction on the Stroop test scores.

**Conclusion:** Complex intersections between ACEs and NLEs impact the phenome of depression, which comprises neuroticism, brooding, suicidal tendencies, and the severity of insomnia, anxiety, and depression, while sexual abuse together with other ACEs and NLEs may impact cognitive interference inhibition.

## Introduction

Depressive episodes are reported by an approximate 4.7% of the worldwide populace within a 12-month timeframe, with the mean age of onset falling within the range of 17 to 37 years.^1^ Depression is a condition that arises from an intricate interaction between genetic, biological, and psychosocial factors.^2^ Depression is distinguished by depressed mood, diminished energy, loss of interest and insufficient daily activity for a minimum of two weeks. These symptoms are often accompanied by additional indications of mental and motor impairment, appetite fluctuations, insomnia, impaired concentration, feelings of worthlessness or unwarranted guilt, and recurrent suicidal ideation.^3,4^

There is substantial evidence to suggest that adverse childhood experiences (ACEs) are associated with a heightened likelihood of detrimental health outcomes, such as suicidal tendencies and depression.^5–9^ ACEs encompass a range of challenges experienced during childhood and adolescence, such as physical, sexual, and emotional abuse; neglect (both physical and emotional); and exposure to dysfunctional households (including but not limited to domestic violence, and parental divorce, substance abuse, mental illness, or criminal behaviors).^10,11^ Neglect and emotional abuse considerably contribute more to the incidence of depression than other types of ACEs, such as sexual abuse and domestic violence, according to a recent meta-analysis.^12^ In contrast, other meta-analysis^13^ discovered a more robust association between depression and sexual and physical abuse compared to the aforementioned categories. Furthermore, ACEs have the potential to modify the default mode network, interfere with reward processing, and reduce the volume of the inferior frontal gyrus. These effects can result in compromised inhibitory control and emotional function.^14–16^ A prospective cohort study discovered cognitive impairments and ACEs that were mediated in part by depressive symptoms.^5^

Additionally, empirical support suggests that daily hassles and negative life events (NLEs) are correlated with symptoms of depression, negative affect, and neuroticism. NLEs and daily difficulties encompass disagreeable and anxiety-inducing obligations such as familial disputes, financial hardships, and work-related obligations.^17,18^ Cognitive dysfunction and emotional dysregulation, as well as an increased risk of developing psychopathology and late-life cognitive impairment, such as major depressive disorder (MDD), may result from daily hassles and exposure to NLEs.^19–25^ MDD was predicted by increased daily hassles in a 14–24-year-old cohort.^23^ Maybery^26^ identified a correlation between perceived distress among university students and adverse interpersonal (e.g., financial, and familial) and non - interpersonal (e.g., academic) events. Longitudinal studies have found that cognitive impairments are significantly predicted by the severity and quantity of daily stressors.^20,21^ It was recently reported that NLEs partially mediate the effects of ACEs on the severity of depression.^27^ This implies that ACEs increase the probability of developing NLEs, which in turn influences the severity of depression. However, the extent to which NLEs partially or completely mediate the effects of ACEs on all features of the phenome of depression (suicidal behaviors, rumination, neuroticism, insomnia, and cognitive deficits) is still unknown.

Consequently, the purpose of this research is to determine whether a) ACEs affect all characteristics of mild MDD in young adults, including suicidal tendencies, brooding, neuroticism, insomnia, cognitive deficits, and the severity of depression and anxiety; and b) whether NLEs partially or wholly mediate these effects. Furthermore, we examine whether solitary ACEs or the combined effects of ACEs are better predictors of the depression phenome. We intend to develop a mediation model that establishes the connection between ACEs and NLEs and depression characteristics, which could be utilized in clinical practice to predict suicidal tendencies and depression. In pursuit of this objective, we model the relationships between all features of the depression phenome using the nomothetic precision psychiatry approach.^27,28^ To assemble a representative sample of individuals diagnosed with mild major depressive disorder (MDD) uncomplicated by chronic depression, we opted to enlist academic students whose mean age corresponds to the onset of depressive symptoms.^1^

## Material and methods

### 2.1. Participants

G*Power 3.1.9.4 was utilized to determine the a priori sample size. The following parameters were established for the case-control study: an analysis of covariance with the following characteristics: f = 0.3 effect size, alpha = 0.05 power, number of groups = 3, and variables = 6. Consequently, a minimum sample size of 111 participants was estimated. To represent young adults, our study invited students of both sexes, aged 18 to 35, from any department of Chulalongkorn University in Bangkok, Thailand, to participate. From November 2021 to February 2023, a cohort of 118 Thai-speaking individuals was enrolled in this research investigation at the outpatient Department of Psychiatry, King Chulalongkorn Memorial Hospital, Bangkok, Thailand. This study enrolled individuals who satisfied the DSM-5-TR^3^ criteria for depressive episode, as diagnosed by senior psychiatrists. Additionally, participants who obtained a Hamilton Depression Rating Scale (HAM-D)^29^ score over 7, as assessed by a clinical psychologist with adequate training, were eligible to participate. For their own protection, participants who exhibited signs of suicidal ideation during the interview phase were ineligible for inclusion in the study and were instead referred to a psychiatrist. The study excluded psychiatric diagnoses such as schizophrenia, alcohol or drug abuse disorders, psycho-organic disorders, anxiety disorders, autism, bipolar disorder, and schizoaffective disorder, in addition to current medical conditions including endocrine or autoimmune disorders, psoriasis, type 1 diabetes, lupus erythematosus, chronic kidney disease, multiple sclerosis, and carcinoma. Female students who were pregnant or nursing were not permitted to take part in the study. Seventy-four patients suffering from an acute phase of major depression were enrolled. Through word-of-mouth and online advertisements, 44 healthy controls devoid of psychiatric diagnoses and suicidality at any point in their lives were recruited for this study. Recruited were controls with a HAM-D^29^ score of 7 or below.

The Institutional Review Board (IRB) of the Faculty of Medicine, Chulalongkorn University, Bangkok, Thailand, reviewed and approved this study (IRB No.351/63). Consent in writing was obtained from every participant.

### 2.2. Methods

The individuals involved in the study were instructed to complete semi-structured interviews regarding sociodemographic information such as age, gender, number of years of education, current smoking status, lifetime history of COVID-19 infection, family history of psychiatric diagnoses (including major depression, bipolar disorder, anxiety, and psychosis), and suicidality. For the computation of body mass index (BMI), weight (kg) and height (m) were acquired. The prevalence of childhood adversities was evaluated for all participants through the utilization of the Adverse Childhood Experiences (ACEs) Questionnaire, as developed by Felitti et al.^10^ Abuse (emotional, physical, and sexual), neglect (emotional and physical), household dysfunction (including domestic violence, substance abuse, mental illness, parental divorce, and criminal members in the household) were the categories applied to a self-report consisting of 28 items. The ACEs Questionnaire underwent a validated Thai translation by Rungmueanporn et al.^30^ The Thai version of the questionnaire demonstrated internal consistency reliability values of 0.79 for the abuse domain, 0.82 for the neglect domain, and 0.66 for the household dysfunction domain.

The Negative Event Scale (NES) was employed to quantify the degree of NLEs.^26^ The self-report questionnaire comprises 42 items, each assigned a score between 0 (indicating the event did not transpire) and 5 (indicating the event transpired with significant problems). The questionnaire assesses four non-interpersonal subscales (health, money, course interest, and academic limitation) and seven interpersonal subscales (friends, boyfriend/girlfriend, parents, relatives, lecturers, and fellow students) that are linked to the current (1-month period) NLEs. The scale, back-translated into Thai by Boonyamalik^31^ demonstrated high reliability among Thai adolescents, as evidenced by Cronbach’s alpha values ranging from 0.94 to 0.96.

The present study utilized the subsequent rating scales to evaluate the phenome of depression. The symptoms and severity of depression were assessed using the Thai version of the HAM-D,^29^ which Lotrakul et al.^32^ translated, and the Beck Depression Inventory-II (BDI-II),^33^ which Mungpanich^34^ translated. The Thai version of the State-Trait Anxiety Inventory (STAI)^35^ was developed by Iamsupasit and Phumivuthisarn.^36^ The STAI comprises twenty items that request respondents to assess their level of state anxiety on a four-point Likert scale ranging from “not at all” (1) to “mostly” (4). The Thai translation of the Insomnia Severity Index (ISI),^37^ a self-rating questionnaire utilized to evaluate the severity of sleep problems, was provided by the Mapi Research Trust.^38^ The ISI^37,38^ is comprised of seven Likert-scale items, where responses span from “no problem” (0) to “very severe problems” (4). The Ruminative Response Scale (RRS),^39^ which was translated into Thai by Thanoi and colleagues,^40^ comprises 22 items utilized to assess reflective rumination and pondering. Responses range from almost never (1) to almost always (4). The total score of eight questions from the Thai version of The International Personality Item Pool-NEO (IPIP-NEO)^41^ was utilized to assess neuroticism, one of the Big Five personality traits, in this research investigation. The Thai version was translated by Yomaboot and Cooper^42^ and utilized items with a Cronbach’s alpha coefficient of 0.83. The researchers utilized the Columbia-Suicide Severity Rating Scale (C-SSRS)^43^ to assess both past (up to one month prior to study enrollment) and present suicidal tendencies (within one month). The severity and intensity of suicidal ideation, the self-harm and attempt subscales, and the lethality subscale comprise the four components of the C-SSRS. The Columbia Lighthouse Project^44^ translated the Thai version of this scale.

Previous studies have established that the traditional Stroop task yielded a significant effect size when used to compare cognitive impairments in patients with clinical depression to those in the control group.^45^ Hence, the present investigation utilized the Stroop Color and Word Test (SCWT) ^46^ to evaluate cognitive interference. The SCWT comprises three stimulus sheets, which feature word, color, and word-color stimuli. Participants are given the task of rapidly identifying the colors of the ink or reading text. Body mass index (BMI) was computed as weight (in kg) divided by length (in meter) squared.

### 2.3 Statistical analyses

The statistical analysis for this study is conducted utilizing IBM, SPSS windows version 29. The researchers utilized a contingency table analysis (Χ^2^-test) to evaluate the statistical associations among categorical variables. Utilizing Pearson’s product-moment correlation coefficients, the statistical relationships between continuous variables were ascertained. Analysis of variance was employed to investigate the relationships between diagnostic categories and clinical data. The researchers utilized analysis of covariance (ANCOVA) to investigate the variation in cognitive variables between groups while covarying for age, sex, and years of education. We used regression analysis to delineate the effects of different ACEs and NLEs on the clinical characteristics of depression. To reduce the number of items into a principal component (PC) score that could be utilized in subsequent statistical investigations, PC analysis (PCA) was implemented. Utilizing Bartlett’s sphericity test and the Kaiser-Meyer-Olkin (KMO) test for sample adequacy, factorability was ascertained; a KMO value greater than 0.6 indicates adequate sample adequacy. Acceptance of the initial PC is contingent upon the variance explained (VE) exceeding 50% and all loadings surpassing 0.66.

Using partial least squares (PLS) path analysis (SmartPLS), ^47^ the causal relationships between latent vectors derived from ACEs, NLEs, cognition, and the depressive phenome were evaluated. PLS path analysis was conducted exclusively on the condition that both the inner and outer models met predetermined quality criteria: a) all loadings on the latent vectors must be greater than 0.66 at a significance level of p<0.001; b) the model fit must be satisfactory when the standardized root mean square residuals (SRMR) do not exceed 0.08; c) all latent vectors must possess adequate composite reliability (>0.7) and Cronbach’s alpha (>0.6) values, with an average variance extracted (AVE) exceeding 0.50; d) the reflective models must not be incorrectly specified. We utilized the Heterotrait-Monotrait ratio (HTMT) with a significance level of 0.85 to assess the discriminant validity of the constructs. A complete PLS analysis was conducted utilizing 5,000 bootstrap samples to ascertain the exact p-values associated with the path coefficients for specific, total indirect, and direct paths.

## Results

### 3.1 Results of PCA

We were unable to extract one validated PC from all ACEs items or subtypes. Nonetheless, we managed to isolate a single PC (referred to as ‘PC ACEs’) from the cumulative score of the emotional abuse, physical abuse, emotional neglect, and physical neglect subcategories, as illustrated in **Table 1**. We constructed four PCs based on specific sets of items: a) The ACEs abuse items 1, 2, 3, and 4, which represent emotional and physical abuse (referred to as ‘PC-ACE abuse’) (KMO=0.776, Bartlett’s sphericity test χ2 =121.660, df=6, p<0.001, VE=57.66%, all loadings>0.740). b) The items 5, 6, and 7, which indicate sexual abuse (referred to as ‘PC-ACE sexual abuse’) (KMO=0.639, Bartlett’s sphericity test χ2 =178.575, df=3, p<0.001, VE=76.14%, all loadings>0.781). c) The items 9, 10, 11, 12, 13, and 15, which represent emotional and physical neglect (referred to as ‘PC-ACE neglect’) (KMO=0.896, Bartlett’s sphericity test χ2 =616.214, df=15, p<0.001, VE=77.44%, all loadings>0.821). d) The ‘ PC-ACE family’, which reflects household dysfunction, including domestic violence, mental illness in the household, and parental divorce subtypes (KMO=0.618, Bartlett’s sphericity test χ2 =23.269, df=3, p<0.001, VE=51.54%, all loadings>0.717). The Electronic Supplementary File (ESF), Table 1 provides a comprehensive explanation of all ACEs items.

**Table 1.** Results of principal component (PC) analyses and construction of adverse childhood experiences (ACEs) and negative life event (NLE) principal components (PCs)

| PC ACEs |  | PC-NLE relationship |  | PC-NLE academic |  | PC Current phenome |  | PC Stroop |  |
| --- | --- | --- | --- | --- | --- | --- | --- | --- | --- |
| Variables | Loading | Variables | Loading | Variables | Loading | Variables | Loading | Variables | Loading |
| Emotional abuse | 0.772 | Hassles with friend | 0.695 | Hassles in academic course | 0.801 | HAM-D | 0.895 | Stroop word | 0.763 |
| Physical abuse | 0.743 | Hassles with parents | 0.722 | Hassles with limitation in academic ability | 0.858 | BDI-II | 0.916 | Stroop color | 0.898 |
| Emotional neglect | 0.807 | Hassles with other students | 0.753 | Hassles in course interest | 0.748 | STAI | 0.781 | Stroop word-color | 0.756 |
| Physical neglect | 0.712 | Hassles with relatives | 0.750 |  |  | ISI | 0.736 |  |  |
|  |  |  |  |  |  | SB | 0.726 |  |  |
|  |  |  |  |  |  | Brooding | 0.894 |  |  |
|  |  |  |  |  |  | Neuroticism | 0.921 |  |  |
| KMO = 0.668 |  | KMO = 0.714 |  | KMO = 0.648 |  | KMO = 0.895 |  | KMO = 0.565 |  |
| X<sup>2</sup> = 129.585<br>df = 6, p <0.001 |  | X<sup>2</sup> = 81.205<br>df = 6, p<0.001 |  | X<sup>2</sup> = 75.079<br>df = 3, p<0.001 |  | X<sup>2</sup> = 680.261<br>df = 21, p<0.001 |  | X<sup>2</sup> = 115.652<br>df = 3, p<0.001 |  |
| VE = 57.655% |  | VE = 53.325% |  | VE = 64.618% |  | VE = 70.942% |  | VE = 68.502% |  |

KMO: The Kaiser-Meyer-Olkin Test; VE: Variance explained; X<sup>2</sup>: Bartlett's test of sphericity. NLEs: negative life events; PC-NLE: principal component of negative life events subcategories (see electronic supplementary file, table1)
NLE\_course: Course, i.e., number of homework/reports/other burdens from your study, the deadline for submitting homework/reports, time pressures, difficulties completing assignments/essays; NLE\_academic limit: Academic limit, i.e., not getting the results you expected, your academic ability not as good as you believe, some subjects are difficult to understand; NLE\_course interest Course interest, i.e., This course is not relevant to your future career, your course is boring.

In addition, we were unable to extract one PC from all NLE items. As a result, we recovered three validated PCs and assigned them the labels ‘PC-NLE self’, ‘PC-NLE relationship’, and ‘PC-NLE academic’, as shown in Table 1 and ESF, Table 1. The total score of all NLEs items was computed (labeled total NLEs).

Based on prior studies,^8,28,48^ we created a reliable and validated psychological construct (PC) to represent the phenome of depression. This construct was developed by considering the combined scores from different measures such as HAM-D, BDI-II, STAI, ISI, neuroticism, brooding, and suicidal behavior (SB).^28^ For more details, please refer to Table 1 and ESF, Table 2. Furthermore, Table 1 presents a PC that was created using three Stroop Color-Word Test (SCWT)^49^ domains: word, color, and word-color. This PC accurately represents the capacity to suppress cognitive interference, which is labelled as PC Stroop.

**Table 2.** Demographic and clinical data of patients with major depression (MDD) with high versus low adverse childhood experiences (ACEs), and healthy controls (HC)

| Variables | HC<br>(n=44) | Low ACEs<br>MDD<br>(n=37) | High ACEs<br>MDD<br>(n=37) | F/$\chi^2$<br>/FET | df | p |
| --- | --- | --- | --- | --- | --- | --- |
| Age (years) | 23.48 (3.18) | 22.79 (3.45) | 21.89 (2.42) | 2.71 | 2/115 | 0.071 |
| Sex (Male/Female) | 7/37 | 5/32 | 7/30 | 0.40 | 2 | 0.818 |
| Education (years) | 16.57 (1.48) | 16.35 (2.87) | 15.19 (2.20) | 4.33 | 2/115 | 0.015 |
| BMI (kg/m<sup>2</sup>) | 22.33 (3.57) | 21.65 (4.42) | 22.81 (5.64) | 0.60 | 2/115 | 0.550 |
| Relationship status<br>(single/relationship) | 27/17 | 17/20 | 13/24 | 5.66 | 2 | 0.064 |
| Current smoking<br>(yes/no) | 1/43 | 4/33 | 2/35 | FET | - | 0.293 |
| Lifetime Covid-19<br>history (yes/no) | 14/30 | 7/30 | 9/28 | 1.80 | 2 | 0.401 |
| Duration of depression<br>(months) | - | 23.24 (20.02) | 25.41 (24.65) | 0.17 | 1/72 | 0.680 |
| **Psychiatric history**<br>**in family** |  |  |  |  |  |  |
| Depression (yes/no) | - | 5/32 | 7/30 | 0.40 | 1 | 0.754 |
| Bipolar disorder<br>(yes/no) | - | 0/37 | 1/36 | FET | - | 1.000 |
| Anxiety disorder<br>(yes/no) | - | 1/36 | 2/35 | FET | - | 1.000 |
| Psychosis (yes/no) | - | 0/37 | 1/36 | FET | - | 1.000 |
| Suicide (yes/no) | - | 3/34 | 4/33 | FET | - | 1.000 |

Results are shown as mean (S.D.); F: results of analysis of variance; $\chi^2$ : analysis of contingency tables; <sup>a</sup>, <sup>b</sup>, <sup>c</sup> pairwise comparisons using LSD (p<0.05), i.e. <sup>a</sup> means significant different from HC, <sup>b</sup> means significant different from Depression-low ACEs, <sup>c</sup> means significant different from Depression-high ACEs.; FET: Fisher-exact probability test.

### 3.2 Socio-demographic and clinical data

**Table 2** displays the socio-demographic and clinical data. The group of patients with MDD was categorized into two subgroups based on their ACEs levels, specifically those with high ACEs and those with low ACEs. This categorization was done using the median-split approach performed on PC ACEs. Depressive patients did not show any significant differences compared to the control group in terms of age, sex, BMI, relationship status, current smoking, or lifetime COVID-19 history. Depressive patients with high ACEs had a downward trend in their level of education compared to other individuals. There were no significant differences observed between patients with high and low ACEs in terms of the length of depression, and family history of mental disorders such as depression, bipolar disorder, anxiety, psychosis, and suicide.

### 3.3 Clinical features of patients with high and low ACEs

**Table 3** provides a comprehensive overview of the clinical characteristics exhibited by both patients and controls. Patients had significantly elevated scores on the HAM-D, BDI-II, STAI, ISI, brooding, neuroticism, present phenome, and SB measures, in comparison to the control group. Patients with elevated ACEs had significantly higher scores on the HAM-D and BDI-II, as well as greater rates of both current and total SB. Additionally, these patients reported a higher prevalence of NLEs in their relationships compared to patients with MDD who had low ACEs scores. The patients had significantly lower Stroop subdomains and cognition scores compared to the controls.

**Table 3.**
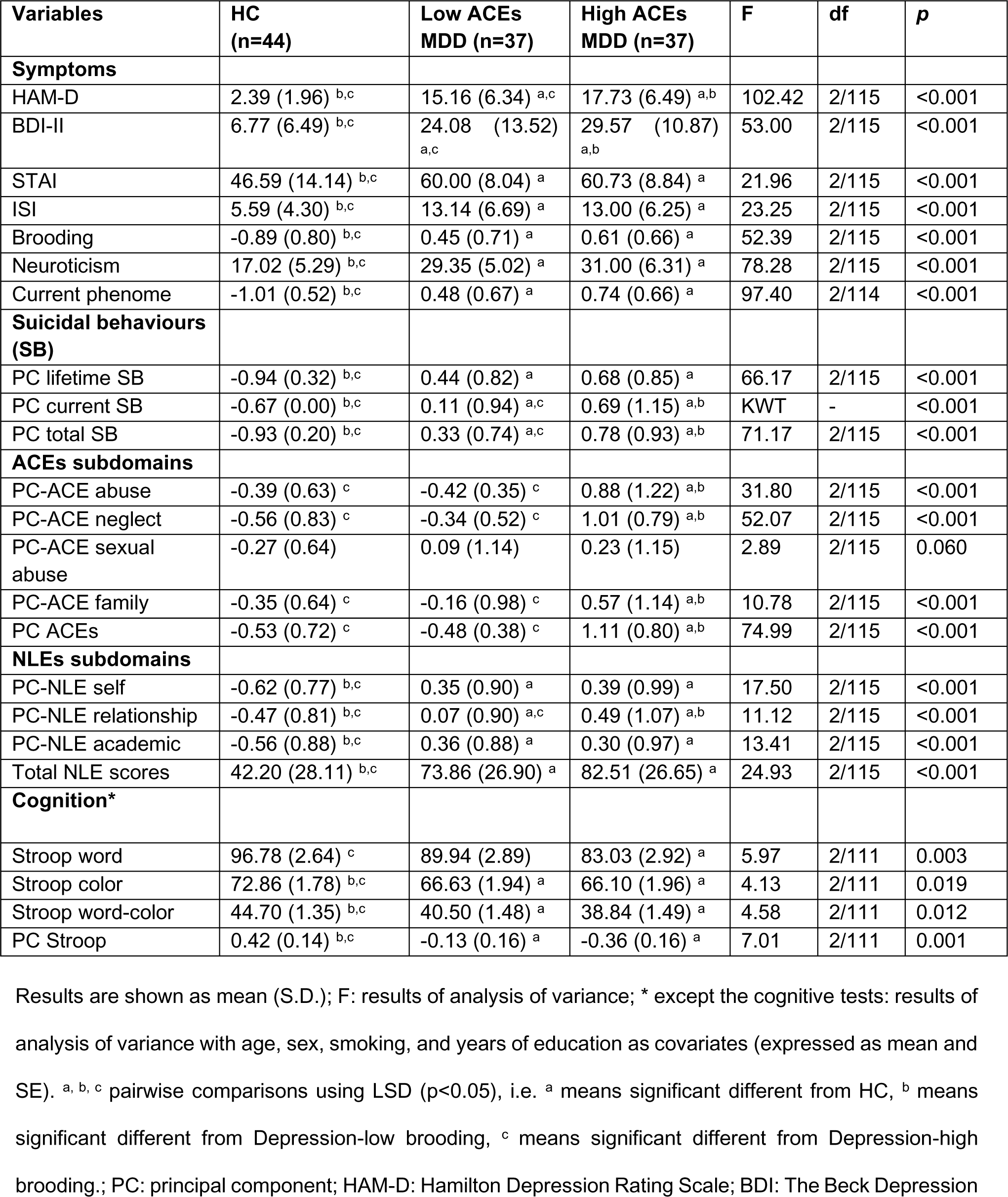
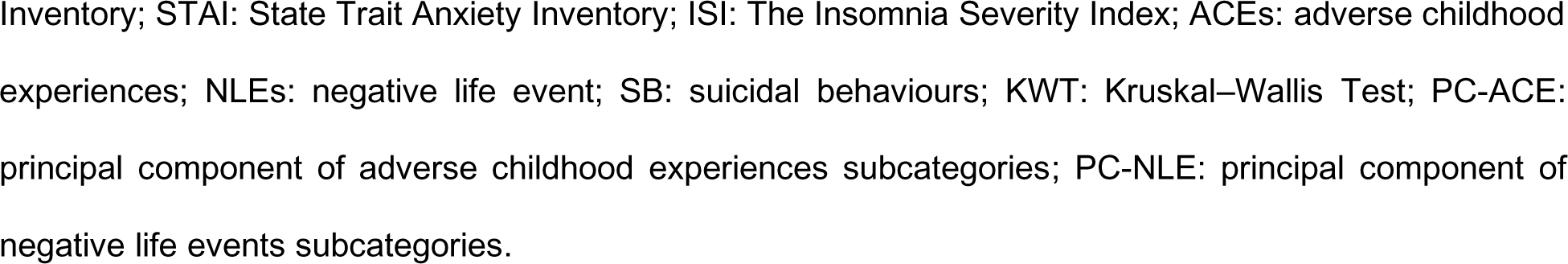
Clinical Features of patients with major depression (MDD) with high versus low adverse childhood experiences (ACEs) and healthy controls (HC)

| Variables | HC<br>(n=44) | Low ACEs<br>MDD (n=37) | High ACEs<br>MDD (n=37) | F | df | p |
| --- | --- | --- | --- | --- | --- | --- |
| **Symptoms** |  |  |  |  |  |  |
| HAM-D | 2.39 (1.96) <sup>b,c</sup> | 15.16 (6.34) <sup>a,c</sup> | 17.73 (6.49) <sup>a,b</sup> | 102.42 | 2/115 | <0.001 |
| BDI-II | 6.77 (6.49) <sup>b,c</sup> | 24.08 (13.52) <sup>a,c</sup> | 29.57 (10.87) <sup>a,b</sup> | 53.00 | 2/115 | <0.001 |
| STAI | 46.59 (14.14) <sup>b,c</sup> | 60.00 (8.04) <sup>a</sup> | 60.73 (8.84) <sup>a</sup> | 21.96 | 2/115 | <0.001 |
| ISI | 5.59 (4.30) <sup>b,c</sup> | 13.14 (6.69) <sup>a</sup> | 13.00 (6.25) <sup>a</sup> | 23.25 | 2/115 | <0.001 |
| Brooding | -0.89 (0.80) <sup>b,c</sup> | 0.45 (0.71) <sup>a</sup> | 0.61 (0.66) <sup>a</sup> | 52.39 | 2/115 | <0.001 |
| Neuroticism | 17.02 (5.29) <sup>b,c</sup> | 29.35 (5.02) <sup>a</sup> | 31.00 (6.31) <sup>a</sup> | 78.28 | 2/115 | <0.001 |
| Current phenome | -1.01 (0.52) <sup>b,c</sup> | 0.48 (0.67) <sup>a</sup> | 0.74 (0.66) <sup>a</sup> | 97.40 | 2/114 | <0.001 |
| **Suicidal behaviours (SB)** |  |  |  |  |  |  |
| PC lifetime SB | -0.94 (0.32) <sup>b,c</sup> | 0.44 (0.82) <sup>a</sup> | 0.68 (0.85) <sup>a</sup> | 66.17 | 2/115 | <0.001 |
| PC current SB | -0.67 (0.00) <sup>b,c</sup> | 0.11 (0.94) <sup>a,c</sup> | 0.69 (1.15) <sup>a,b</sup> | KWT | - | <0.001 |
| PC total SB | -0.93 (0.20) <sup>b,c</sup> | 0.33 (0.74) <sup>a,c</sup> | 0.78 (0.93) <sup>a,b</sup> | 71.17 | 2/115 | <0.001 |
| **ACEs subdomains** |  |  |  |  |  |  |
| PC-ACE abuse | -0.39 (0.63) <sup>c</sup> | -0.42 (0.35) <sup>c</sup> | 0.88 (1.22) <sup>a,b</sup> | 31.80 | 2/115 | <0.001 |
| PC-ACE neglect | -0.56 (0.83) <sup>c</sup> | -0.34 (0.52) <sup>c</sup> | 1.01 (0.79) <sup>a,b</sup> | 52.07 | 2/115 | <0.001 |
| PC-ACE sexual abuse | -0.27 (0.64) | 0.09 (1.14) | 0.23 (1.15) | 2.89 | 2/115 | 0.060 |
| PC-ACE family | -0.35 (0.64) <sup>c</sup> | -0.16 (0.98) <sup>c</sup> | 0.57 (1.14) <sup>a,b</sup> | 10.78 | 2/115 | <0.001 |
| PC ACEs | -0.53 (0.72) <sup>c</sup> | -0.48 (0.38) <sup>c</sup> | 1.11 (0.80) <sup>a,b</sup> | 74.99 | 2/115 | <0.001 |
| **NLEs subdomains** |  |  |  |  |  |  |
| PC-NLE self | -0.62 (0.77) <sup>b,c</sup> | 0.35 (0.90) <sup>a</sup> | 0.39 (0.99) <sup>a</sup> | 17.50 | 2/115 | <0.001 |
| PC-NLE relationship | -0.47 (0.81) <sup>b,c</sup> | 0.07 (0.90) <sup>a,c</sup> | 0.49 (1.07) <sup>a,b</sup> | 11.12 | 2/115 | <0.001 |
| PC-NLE academic | -0.56 (0.88) <sup>b,c</sup> | 0.36 (0.88) <sup>a</sup> | 0.30 (0.97) <sup>a</sup> | 13.41 | 2/115 | <0.001 |
| Total NLE scores | 42.20 (28.11) <sup>b,c</sup> | 73.86 (26.90) <sup>a</sup> | 82.51 (26.65) <sup>a</sup> | 24.93 | 2/115 | <0.001 |
| **Cognition\*** |  |  |  |  |  |  |
| Stroop word | 96.78 (2.64) <sup>c</sup> | 89.94 (2.89) | 83.03 (2.92) <sup>a</sup> | 5.97 | 2/111 | 0.003 |
| Stroop color | 72.86 (1.78) <sup>b,c</sup> | 66.63 (1.94) <sup>a</sup> | 66.10 (1.96) <sup>a</sup> | 4.13 | 2/111 | 0.019 |
| Stroop word-color | 44.70 (1.35) <sup>b,c</sup> | 40.50 (1.48) <sup>a</sup> | 38.84 (1.49) <sup>a</sup> | 4.58 | 2/111 | 0.012 |
| PC Stroop | 0.42 (0.14) <sup>b,c</sup> | -0.13 (0.16) <sup>a</sup> | -0.36 (0.16) <sup>a</sup> | 7.01 | 2/111 | 0.001 |

Results are shown as mean (S.D.); F: results of analysis of variance; \* except the cognitive tests: results of analysis of variance with age, sex, smoking, and years of education as covariates (expressed as mean and SE). <sup>a, b, c</sup> pairwise comparisons using LSD ( $p < 0.05$ ), i.e. <sup>a</sup> means significant different from HC, <sup>b</sup> means significant different from Depression-low brooding, <sup>c</sup> means significant different from Depression-high brooding.; PC: principal component; HAM-D: Hamilton Depression Rating Scale; BDI: The Beck Depression
Inventory; STAI: State Trait Anxiety Inventory; ISI: The Insomnia Severity Index; ACEs: adverse childhood experiences; NLEs: negative life event; SB: suicidal behaviours; KWT: Kruskal–Wallis Test; PC-ACE: principal component of adverse childhood experiences subcategories; PC-NLE: principal component of negative life events subcategories.

### 3.4 Intercorrelation matrix

**Table 4** displays the associations between PC ACEs, PC-ACE sexual abuse and PC-ACE family, as well as total scores of NLEs, and various clinical features of depression. These features include HAM-D, BDI-II, STAI, ISI, brooding, neuroticism, present phenome, lifetime and current SB, and cognition. Positive correlations were found between PC ACEs and total NLEs with all clinical characteristics, but negative associations were observed with cognition. The PC-ACE sexual abuse showed a strong negative correlation with cognitive abilities, while it exhibited a positive correlation with the total scores of NLEs, ISI, and lifetime SB. PC ACEs, Total NLEs, and PC-ACE family (excluding cognition) were strongly linked to every aspect of depression.

**Table 4.** Correlation matrix between adverse childhood experiences (ACEs) and negative life events (NLEs) and other clinical features of depression.

| Variables | PC ACEs | PC-ACE sexual abuse | PC-ACE family | NLEs |
| --- | --- | --- | --- | --- |
| PC-ACEs sexual abuse | 0.285** | - | - | - |
| PC-ACEs family | 0.387*** | 0.152 | - | - |
| NLEs | 0.440*** | 0.353*** | 0.343*** | - |
| HAM-D | 0.424*** | 0.058 | 0.301*** | 0.592*** |
| BDI-II | 0.436*** | 0.031 | 0.326*** | 0.628*** |
| STAI | 0.251** | 0.118 | 0.211* | 0.554*** |
| ISI | 0.300*** | 0.195* | 0.272** | 0.491*** |
| Brooding | 0.355*** | 0.133 | 0.232* | 0.599*** |
| Neuroticism | 0.376*** | 0.103 | 0.200* | 0.550*** |
| PC current phenome | 0.438*** | 0.111 | 0.280** | 0.646*** |
| PC lifetime SB | 0.462*** | 0.233* | 0.324*** | 0.533*** |
| PC current SB | 0.358*** | -0.023 | 0.293** | 0.485*** |
| PC total SB | 0.473*** | 0.127 | 0.344*** | 0.553*** |
| PC Stroop | -0.248** | -0.323*** | -0.084 | -0.219* |

We conducted multiple regression analyses to examine the relationship between the current phenome, including current SB, and cognitive impairments. We included age, sex, number of education years, current regular smoking, and PCs of ACEs and NLEs subtypes as explanatory factors. The results are presented in **Table 5**. The stepwise technique revealed that the current phenome (#regression 1) was most accurately predicted by PC-NLE self, PC-NLE academic, and PC-ACE abuse, which collectively accounted for 53.4% of its variability. The most accurate predictors of current SB were PC-ACE neglect and PC-ACE family, which accounted for 17.8% of the observed variation in the data (regression 2). In Regression #3, specifically, the predictor variable of PC-NLE relationship accounted for 15.5% of the variance in current SB. The cognitive PC (#regression 4) was most accurately predicted by PC-ACE sexual abuse, PC-ACE neglect, and current smoking, which collectively accounted for 18.5% of the variability. **Figure 1** displays the partial regression analysis of the present phenome on PC-NLE self, while accounting for the influences of age, sex, number of schooling years, and current smoking. **Figure 2** displays the partial regression analysis of current SB on PC-ACE neglect, while accounting for the influence of age, sex, number of schooling years, and current smoking.

**Figure 1.**
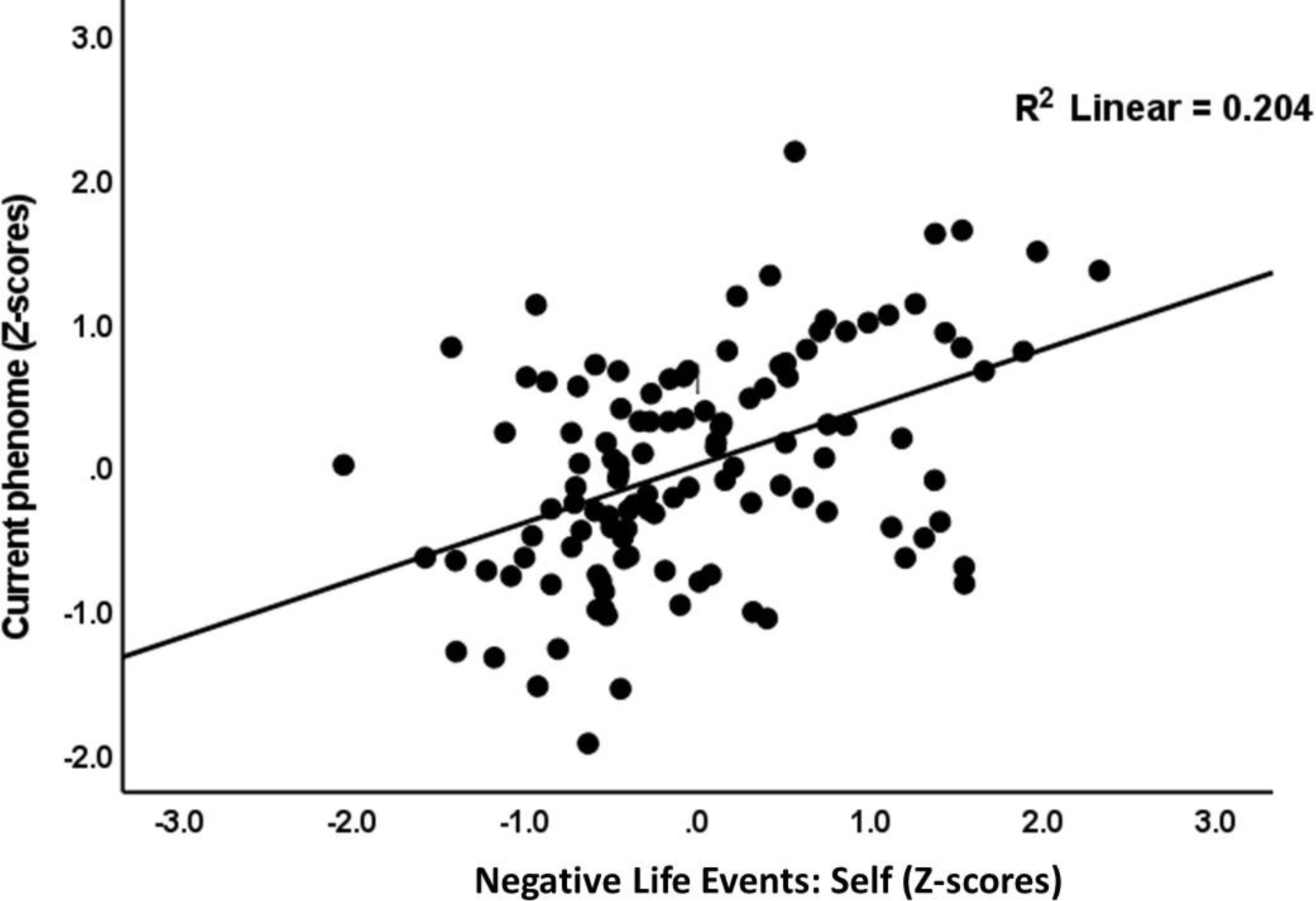
Partial regression of the current phenome score on principal component of negative life events in self-subcategories (after controlling for the effects of age, sex, number of education years, and current smoking).

**Figure 2.**
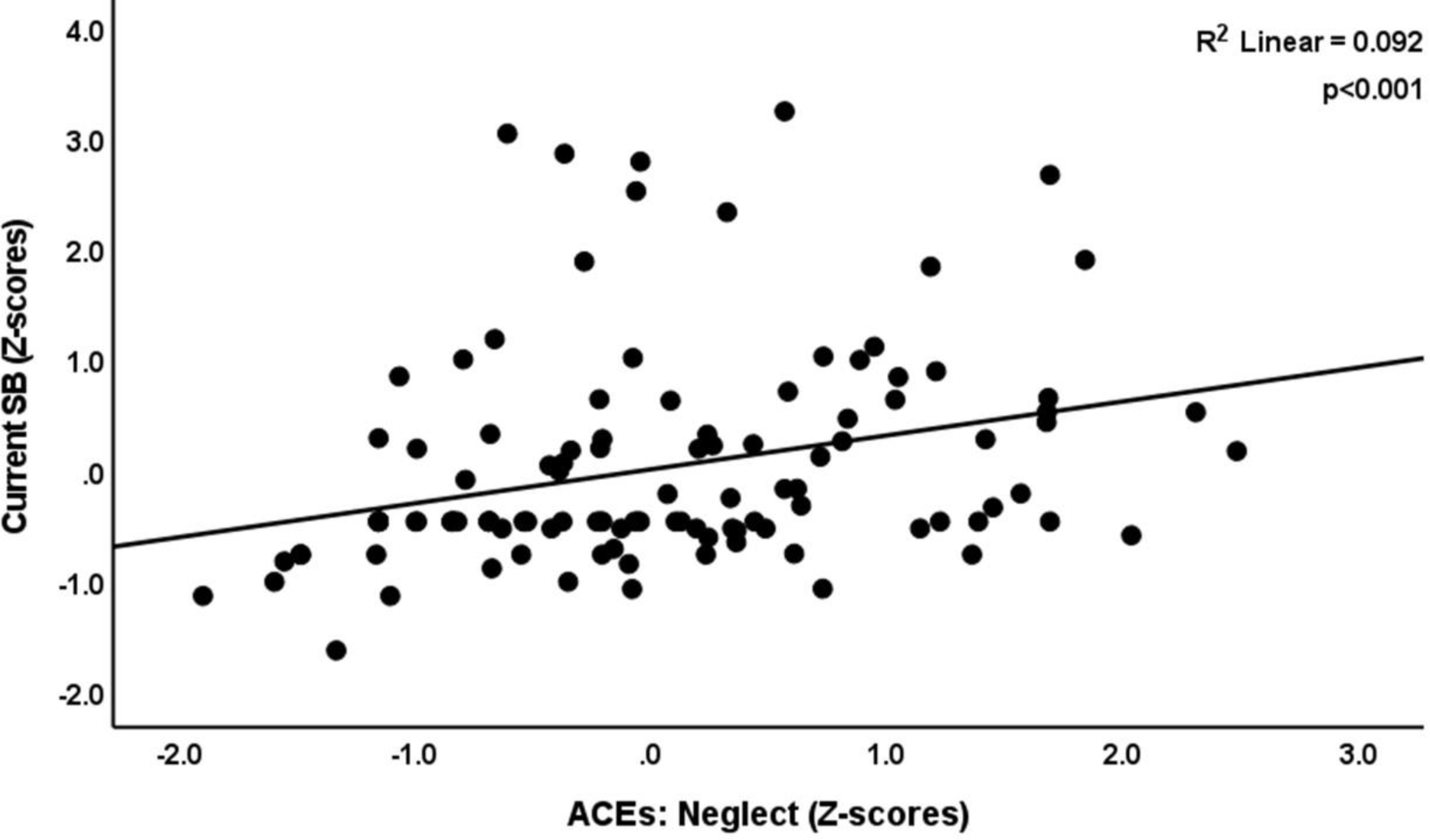
Partial regression of current suicide behaviors (SB) on principal component of adverse childhood experiences in neglect-subcategories (after controlling for the effects of age, sex, number of education years, and current smoking).

**Table 5.** Results of multiple regression analyses with current phenome, suicidal behaviors and neurocognition as dependent variables and adverse childhood experiences (ACEs) and negative life events (NLEs) as input variables.

| Dependent variables | Explanatory variables | Parameter estimates + statistics |  |  | Model statistics + effect size |  |  |  |
| --- | --- | --- | --- | --- | --- | --- | --- | --- |
| | | $\beta$ | t | p | F model | df | p | R<sup>2</sup> |
| #1 PC Current phenome | Model |  |  |  | 43.195 | 3/113 | <0.001 | 0.534 |
|  | PC-NLE self | 0.403 | 5.381 | <0.001 |  |  |  |  |
|  | PC-NLE academic | 0.346 | 4.643 | <0.001 |  |  |  |  |
|  | PC-ACE abuse | 0.267 | 4.130 | <0.001 |  |  |  |  |
| #2 PC Current SB | Model |  |  |  | 11.784 | 2/115 | <0.001 | 0.170 |
|  | PC-ACE neglect | 0.307 | 3.421 | <0.001 |  |  |  |  |
|  | PC-ACE family | 0.193 | 2.151 | 0.034 |  |  |  |  |
| #3 PC Current SB | Model |  |  |  | 21.226 | 1/116 | <0.001 | 0.155 |
|  | PC-NLEs relationship | 0.393 | 4.607 | <0.001 |  |  |  |  |
| #4 PC Stroop | Model |  |  |  | 8.628 | 3/114 | <0.001 | 0.185 |
|  | PC-ACE sexual abuse | 0.279 | 3.218 | 0.002 |  |  |  |  |
|  | PC-ACE neglect | 0.224 | 2.583 | 0.011 |  |  |  |  |
|  | Current smoking | 0.193 | 2.280 | 0.024 |  |  |  |  |

'PC-ACEs abuse' reflects the emotional abuse and physical abuse subcategories of ACEs; 'PC-ACEs neglect' reflects the emotional neglect and physical neglect subcategories of ACEs; 'PC-ACEs sexual abuse' reflects sexual abuse subcategories of ACEs; 'PC-ACEs family' reflects household dysfunction, including domestic violence, mental illness in the household, and parental divorce subcategories of ACEs; SB: suicidal behaviours; PC-ACE: principal component of adverse childhood experiences subcategories; PC-NLE: principal component of negative life events subcategories.

### 3.5 Results from PLS analysis

**Figure 3** displays the final PLS model, highlighting the significant paths. Based on prior research,^8,28,48^ we create a latent vector representing the phenome of depression using the HAM-D, BDI-II, STAI, ISI, brooding, neuroticism, and current SB scores. We successfully developed a factor by considering ACEs abuse, neglect, and familial dysfunctions. We created a validated latent vector for NLEs by utilizing self, relationships, and academic scores. The three Stroop domains were utilized to form a cognitive factor. We utilized the ACEs and NLEs factors and sexual abuse as indicators for predicting the depression phenome and cognitive functioning. Additionally, we examined how NLEs can work as mediators for the impact of ACEs on the phenome. In addition, we incorporated a moderator model that encompasses the ACEs factor and the impact of NLEs on the phenome. The model fit indicates a satisfactory level of quality, as evidenced by the SRMR value of 0.058. The ACE s, NLEs, current phenome, and cognition factors demonstrate sufficient construct validity and convergence. The AVEs for these factors are 0.560, 0.629, 0.708, and 0.684, respectively. The Cronbach’s alpha values for these factors are 0.610, 0.704, 0.929, and 0.766, respectively. Additionally, the composite reliability (rho_C) values for these factors are 0.791, 0.835, 0.944, and 0.866, respectively. All loadings of the outer model exceeded 0.7, except for family difficulties, which had a loading of 0.692. The ACEs and NLEs factors accounted for 55.0% of the phenome variance, while the ACE indicators explained 22.5% of the NLEs variance. Sexual abuse and the phenome factor accounted for 18.5% of the variance in the cognition factor. The moderation term “ACEs X NLEs” was significant. The ACEs factor had a significant specific indirect effect on cognition, which was mediated via the pathway from the NLEs factor to the phenome factor (t=2.38, p=0.017). Furthermore, the NLEs factor had a specific indirect impact on the cognition factor mediated by the phenome, as evidenced by a statistically significant t-value of 2.88 and a p-value of 0.004. The PLS Predict analysis indicated that all Q^2^ values were above zero. Additionally, the PLS cross-validated predictive ability test analysis revealed that the PLS-SEM against indicator average had a significant result (p=0.008), suggesting that the model is replicable.

**Figure 3.**
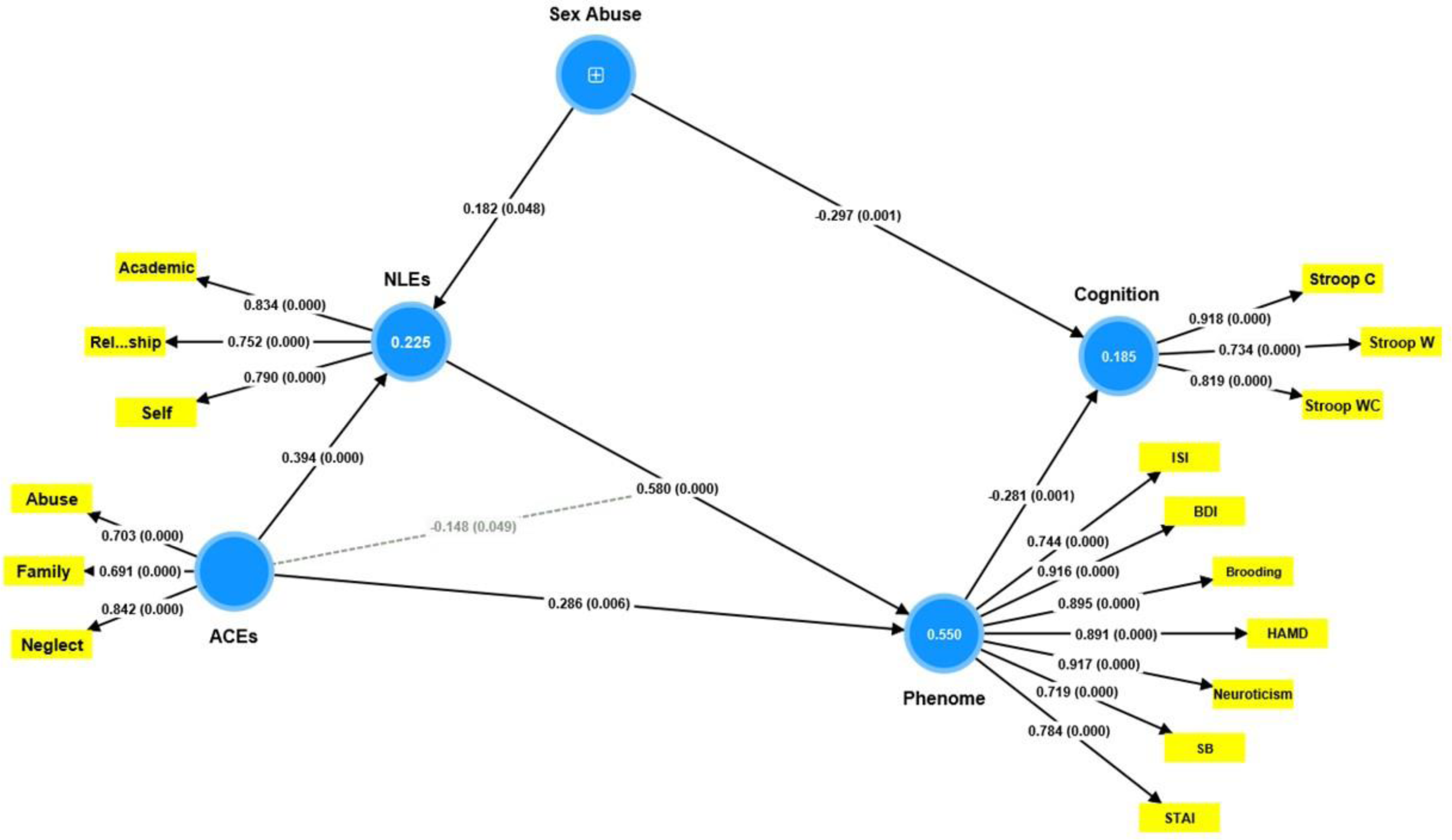
Results of PLS analysis. The significant pathways are presented, including the path coefficients (along with their exact p values) of the inner model and the loadings (with p values) of the outer model. One factor was extracted from the subcategories in adverse childhood experiences (ACEs), such as abuse, neglect, and family dysfunction. Sexual abuse was included as a single indicator (denoted as +). Negative life events (NLEs), the depression phenome, and cognition were entered as latent vectors extracted from their manifestations. Figures in blue circles indicate explained variances. Shown are path coefficients (with p values) and factor loadings (with p values). ‘Abuse’ reflects the emotional and physical abuse subcategories of ACEs; ‘Neglect’ reflects the emotional and physical neglect subcategories of ACEs; ‘Family’ reflects household dysfunction, including domestic violence, mental illness in the household, and parental divorce subcategories of ACEs; ‘Self’ reflects NLEs self-subcategories, such as hassles in health issues and insufficient money; ‘Academic’ reflects NLEs academic-subcategories, such as hassles in academic course, course interest, and limitation in academic ability; ‘Relationship’ reflects NLEs relationship-subcategories, such as hassles with friends, parents, relatives, and other students; HAM-D: Hamilton Depression Rating Scale; BDI-II: The Beck Depression Inventory; STAI: State Trait Anxiety Assessment; ISI: The Insomnia Severity Index; SB: suicidal behaviors. Stroop W: Stroop word; Stroop C: Stroop color; Stroop WC: Stroop word-color.

## Discussion

### 4.1 Effects of ACEs on the depression phenome

The first important finding from this research is that ACEs were substantially correlated with the phenome of depression. Our research distinguished the various ACEs types using a data-centric approach and identified the significant pathways by which ACEs may influence the phenome via PLS analysis. A factor extracted from physical and emotional maltreatment and neglect as well as family problems had a substantial, significant direct effect on the phenome, according to this analysis. These results build upon the conclusions drawn by Almulla et al.,^22^ which indicated that ACEs could predict the phenome of depression. Consistent with previous research (e.g., Hein et al.;^50^ Peckins et al.;^51^ Gizdic et al.^52^), our findings demonstrate that distinct subtypes of ACEs are associated with psychopathology in unique ways. Prior studies have established a correlation between neglect, physical and emotional maltreatment, and an elevated susceptibility to psychological and behavioral complications, including depression and suicidal ideation.^53^ Similarly, an increase in depressive symptoms is correlated with higher cumulative ACEs scores.^54^ The threat-deprivation model, alternatively referred to as the Dimension Model of Adversity and Psychopathology, elucidates the biological mechanisms that underlie a multitude of ACEs.^14,55^ Threat is defined as initial exposure to violence or injury, while deprivation is characterized by the absence of anticipated demands from one’s surroundings.^14,55^ There is evidence that individuals who are threatened or deprived may display increased emotional reactivity and struggle to regulate their emotions.^56^ This has been associated with symptoms of anxiety and depression.^57,58^ Moreover, there is a positive correlation between physical abuse history and poverty among adults^59^ which is linked to depression and anxiety.^60^ According to Dube et al.,^11^ ACEs are strongly associated with suicide attempts.

Furthermore, by incorporating the ACEs items pertaining to parental divorce, domestic violence, mental illness, and suicide within the household, our research successfully constructed a validated family dysfunction factor. A potential explanation for the high loadings of those different items on the latter factor is that all items could be attributed to hereditary influences. This factor exhibits a significant association with every characteristic of depression, except for cognitive dysfunctions. MDD patients exhibit heritability ranging from 30% to 50% explained variance, suggesting that genetics significantly impact the development of depression.^61^ Furthermore, there is evidence to suggest that parental mental health issues account for around 50% of the genetic transmission of suicide attempts.^62^ Furthermore, completed suicide has been linked to genetic factors.^63^ Therefore, it is probable that the existence of mental disorders, regardless of their genetic predisposition, contributed to domestic violence and parental divorce, as well as to the exacerbation of depression symptoms such as suicidal ideation, brooding, insomnia, and overall severity of the condition.

### 4.2. Effects of sexual abuse on the depression phenome

The results of our study indicate that sexual abuse did not have a statistically significant direct impact on the phenome of depression. However, it did have a minimal effect size on lifetime SB and ISI scores. These results are consistent with those of Gizdic et al.,^52^ who discovered no association between sexual abuse and psychopathology using PCA. However, our research revealed that sexual abuse exerted a substantial indirect influence on the phenome of depression, and that this influence was mediated by NLEs. Positive results indicating a significant association between childhood sexual abuse and depression are also explicable by this effect. As an illustration, a longitudinal investigation conducted by Ferguson et al.^64^ identified a correlation that was statistically significant between sexual abuse and various socioecological outcomes pertaining to adult development, including psychological well-being, sexual risk-taking, and physical health. Sexual abuse was statistically associated with a lifetime diagnosis of psychiatric disorders, including anxiety disorder, depression, eating disorders, posttraumatic stress disorder, sleep disorders, and suicide attempts, according to meta-analysis and longitudinal study.^65,66^ A higher probability of exhibiting symptoms consistent with severe depression was found in individuals whose sexual abuse took place prior to the age of twelve, according to a study by Schoedl et al.^67^ Adams et al.^68^ discovered that, unlike during childhood and adolescence, a prior history of sexual abuse prior to the age of six did not exhibit a significant predictive effect on psychopathology.

### 4.3 Combined effects of ACEs and NLEs on the phenome of depression

A third significant finding of the present study is that ACEs and NLEs are predictors of the MDD phenotype, which includes suicidal behaviors. Self- and academic-related difficulties in conjunction with ACEs predicted the depression phenome, whereas neglect, family dysfunction, or relationship difficulties predicted current suicidal behaviors. Chronic stress can be induced by everyday difficulties, particularly when these difficulties coincide with other personal and environmental risk factors that increase the likelihood of developing mental disorders.^23^ Previous studies have established clear associations between NLEs and neuroticism,^69^ brooding,^70,71^ poor sleep quality,^70,72^ suicide,^27,71^ and all of these factors are known to increase the likelihood of developing depression.^23,24,73^

Furthermore, increased NLEs were found to partially mediate the effects of ACEs on rumination, SB, neuroticism, and the phenome of depression, according to the current study. Almulla et al. ^22^ discovered that NLEs combined with ACEs substantially predict the manifestation of very severe MDD, as well as the lifetime and current suicidal behaviors. These results build upon those of Almulla et al.^22^ The effects of ACEs on the increases in affective symptoms during the COVID-19 pandemic are partially mediated by increased NLE scores.^74^ Maes and Almulla^27^ identified a similar pattern to the present study: ACEs predict the severity of NLEs, and the latter are correlated with heightened illness severity and recent suicidal behaviors. As previously mentioned, individuals who have experienced ACEs are more likely to reside in environments that are more problematic, which increases their exposure to NLEs.^27^

Furthermore, we identified a noteworthy moderating effect in which dysfunctions within the family and physical and emotional neglect and abuse serve as moderators to diminish the impact of NLEs on the phenome of depression. Another study that documented this moderation effect was Maes and Almulla.^27^

### 4.4 ACEs predict cognitive impairments in MDD

The fourth significant discovery of this research is that ACEs are notably correlated with cognitive impairments in individuals with depression. Significantly, our research revealed that various ACEs had distinct impacts on cognitive impairments associated with depression. Specifically, sexual abuse had direct effects, while neglect and family issues and a combination of physical and emotional abuse had indirect effects. The latter effects were mediated in part by NLEs and the depressive phenome.

The Classic Stroop test was employed in this investigation as it was deemed a reliable benchmark for depression in a prior meta-analysis.^45^ Cognitive interference occurs when the processing of one attribute influences the simultaneous processing of another attribute in response to the same stimuli.^46,75^ The Stroop test measures the capacity to prevent cognitive interference. Interference control is a critical cognitive principle that is potentially indispensable for numerous memory tasks and exhibits connections with other cognitive processes.^76^ Like working memory and set shifting (flexibility), inhibitory control is an essential component of the executive functions, which are higher-order cognitive processes.^77^ Thus, our findings extend those of Tjoelker et al.,^78^ who found that a history of physical maltreatment and emotional neglect is associated with diminished interference control in older individuals with depressive, anxious, and somatic symptoms. A significant correlation was identified by Lund et al. ^79^ in a systematic review between ACEs and executive deficits in children, specifically in relation to maternal depression. According to a study by Saleh et al.,^80^ exposure to ACEs negatively impacted working memory performance and processing speed. In a study of adults aged 18-45 years, both with and without MDD, who had been exposed to ACEs before the age of 13, Gould et al.^81^ found that ACEs may cause impairments in affective processing/inhibition, spatial working memory, and executive functions. Furthermore, ACEs have the potential to induce cognitive dysfunctions, disrupt reward systems, promote avoidant-emotion coping, and reduce problem-focused coping strategies. These combined effects may heighten an individual’s susceptibility to NLEs.^11,14,82^

Significantly, our PLS analysis revealed that sexual abuse exerted a considerable direct impact on cognitive impairments. These results build upon the research conducted by Barrera et al.,^83^ which suggested that children who have been subjected to sexual assault might struggle to maintain focus. Further supporting the results are those of Hawkins et al.,^84^ who found that neglect and sexual abuse were associated with decreased performance on short-term and long-term memory assessments among individuals aged 24-32.

### 4.5. Different pathways may explain the effects of ACEs/NLEs on the phenome of depression

The impact of physical and emotional abuse and neglect on the manifestation of depression could potentially be elucidated through the influence of ACEs on various cognitive function-affecting pathways. ACEs may increase the production of M1 macrophages and T-helper (Th) cells, specifically Th-1 and Th-17 cells, thereby activating the immune-inflammatory response system (IRS), while diminishing neuroprotective factors such as nerve growth factor.^9,85^ Furthermore, nitro-oxidative pathways are stimulated, and antioxidant defenses are compromised by ACEs.^86–88^ Furthermore, research suggests that ACEs, such as neglect, could potentially interfere with the functioning of the hypothalamic-pituitary-adrenal (HPA) axis.^89,90^ Fourthly, childhood exposure to domestic violence shortened telomere length, according to a meta-analysis^91^ and ACEs can alter epigenetic methylation patterns of HPA-axis-related genes.^92^ Furthermore, it has been observed that ACEs are linked to alterations in the microbiome, which result in a distinct enterotype reflecting dysbiosis of the gut.^7^ ACEs elevate atherogenicity, a factor that has been linked to suicidal tendencies and depression.^93^ Finally, ACEs have been linked to reduced volume in the amygdala, hippocampus, and anterior cingulate cortex—regions of the brain responsible for emotion regulation and self-control—along with impairments in executive functions, which are indicative of impaired memory and cognitive flexibility.^6,25,84,94–96^

The potential impact of sexual abuse on neurocognitive function could be accounted for by the known cognitive function-altering effects of ACEs on various pathways, such as inflammation,^66,88,97^ alterations in the HPA-axis,^97^ and disruptions in the cortical and subcortical regions.^98^ Likewise, NLEs have a plethora of detrimental effects on the same pathways, including the IRS,^99,100^ oxidative stress pathways,^101^ HPA-axis function,^102^ and methylation processes.^102^

### 4.5 Limitations

Our research was limited to depression among university students between the ages of 18 and 35; consequently, the findings might not be applicable to chronic and recurrent depression, and depression in old age. Furthermore, the age of initial exposure to ACEs was not included in the Thai version of the Adverse Childhood Experiences Questionnaire,^10,30^ which was utilized in this research. Third, recall bias may be present due to the retrospective nature of the ACEs assessment in this study.^103^ While some may contend that a more substantial sample size is imperative for examining the associations between ACEs/NLEs and the phenome, our research utilized a sample size that possessed an adequate power of 0.8. Furthermore, the association between ACEs/NLEs and the phenome has a post-hoc power of 1.0. Furthermore, the model exhibited replicability and the fit quality of the data was satisfactory. Further investigation into associated biomarkers is necessary to establish a more precise intervention strategy, given the involvement of ACEs in neurobiological pathways.

#### Conclusions

A significant proportion of the variability observed in the phenome of depression was accounted for by the combined influence of ACEs, including family dysfunction, abuse, and neglect (both physical and emotional), and self-, relationship, and academic-related NLEs. ACEs interacted substantially with NLEs to influence the depression phenome. While sexual abuse did not exert a direct influence on the phenome, its consequences were mediated through NLEs. We discovered that increased sexual abuse, physical and emotional abuse and neglect, and family dysfunction-related adverse childhood experiences predicted 22.5% of the variance in NLEs. Sexual abuse and depression as a phenotype accounted for as much as 18.5% of the variance in Stroop test scores. Sexual abuse coupled with other ACEs and NLEs may have an effect on cognitive interference inhibition.

## Acknowledgements

None

## Author contributions

AV and MM designed the current study and performed the statistical evaluation. AV contributed to the data collection. All authors contributed to the writing and rewriting of the manuscript, and they have all given their consent for submission of the completed version.

## Ethics statement

The studies involving humans were approved by The Institutional Review Board of Chulalongkorn University’s institutional ethics board, Bangkok, Thailand. The studies were conducted in accordance with the local legislation and institutional requirements. The participants provided their written informed consent to participate in this study. Written informed consent was obtained from the individual(s) for the publication of any potentially identifiable images or data included in this article.

## Data availability statement

The raw data supporting the conclusions of this article will be made available by the authors, without undue reservation.

## Electronic Supplementary File (ESF)

**ESF, Table 1.**
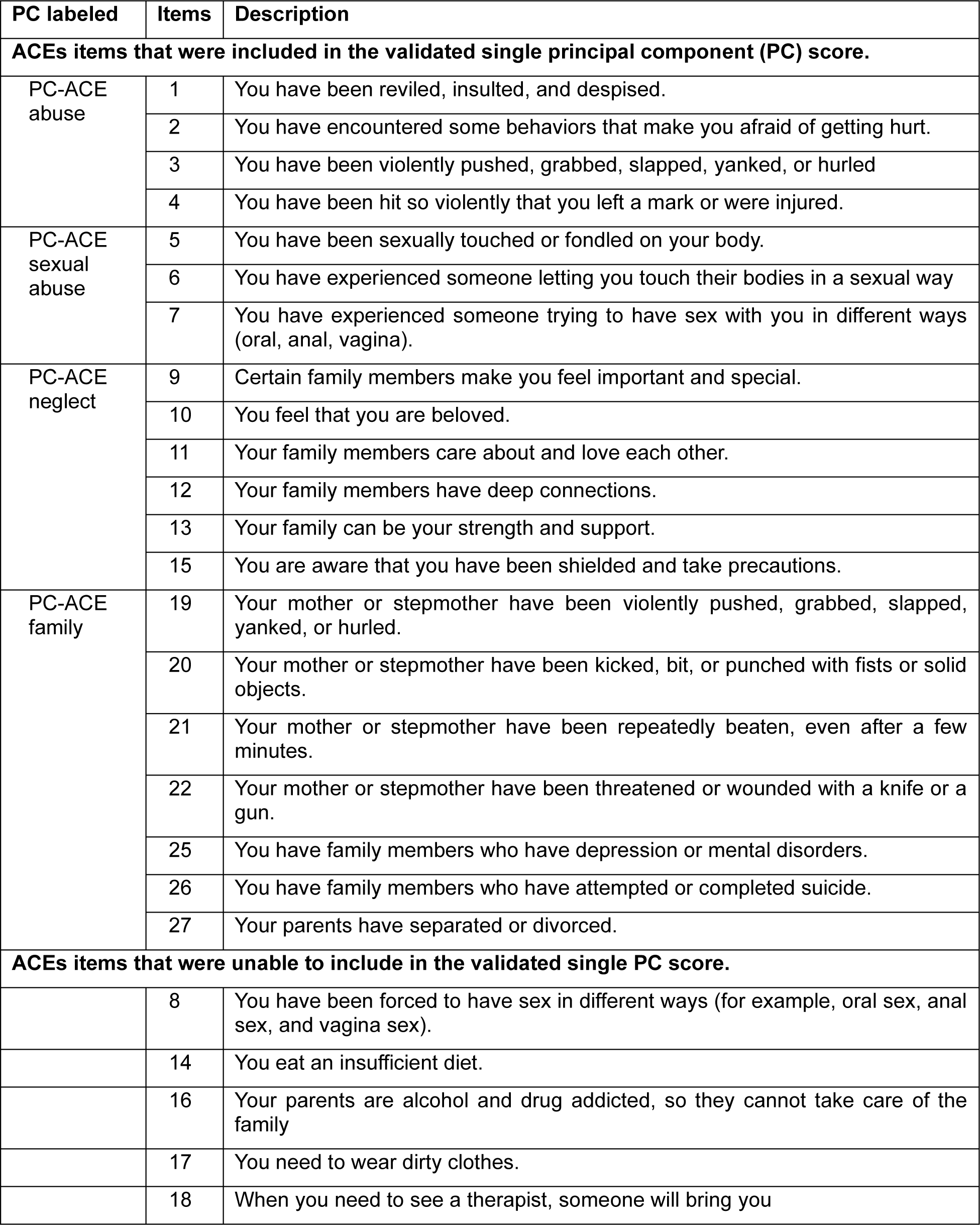
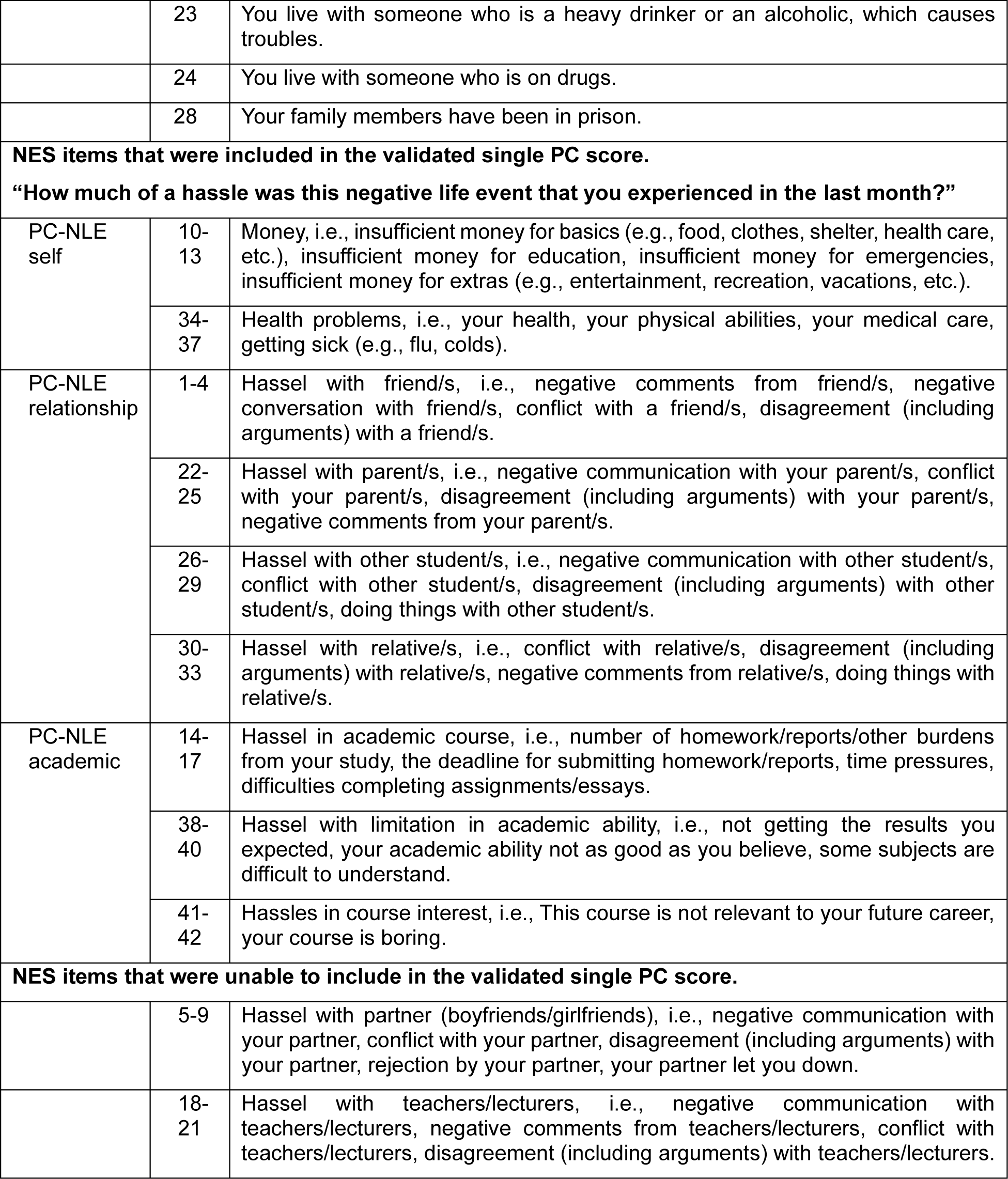
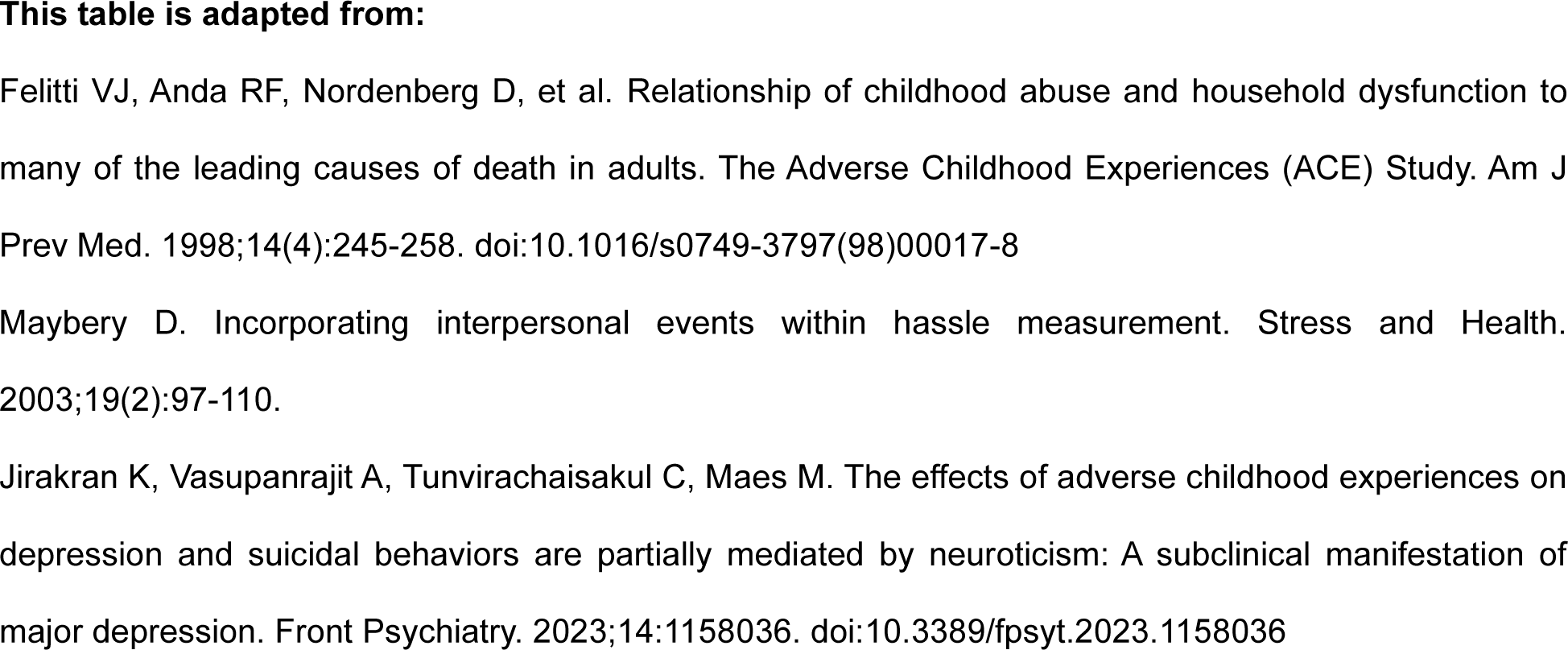
Description of childhood adversities from The Adverse Childhood Experiences (ACEs) Questionnaire and negative life events (NLEs) from The Negative Event Scale (NES).

**ESF, Table 2.**
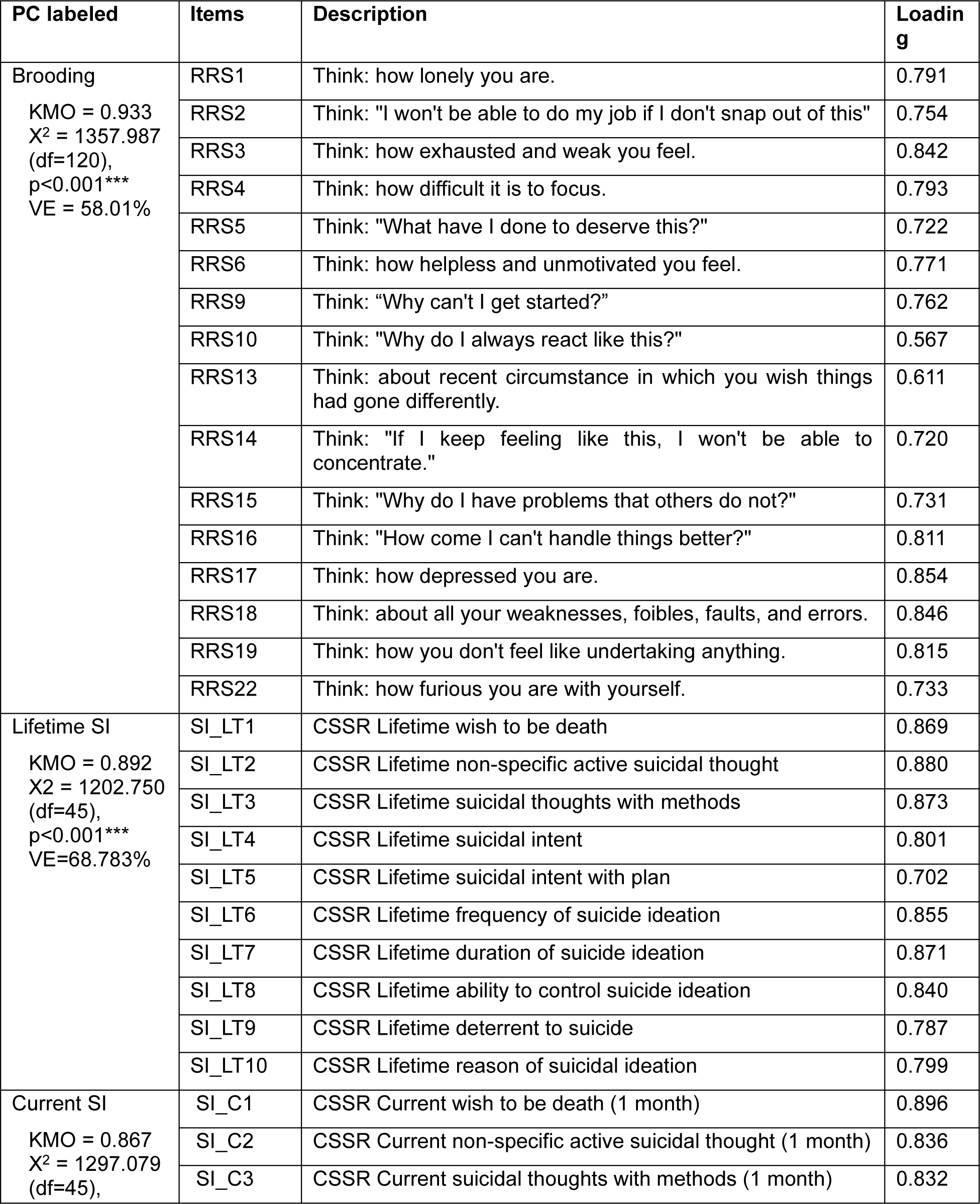
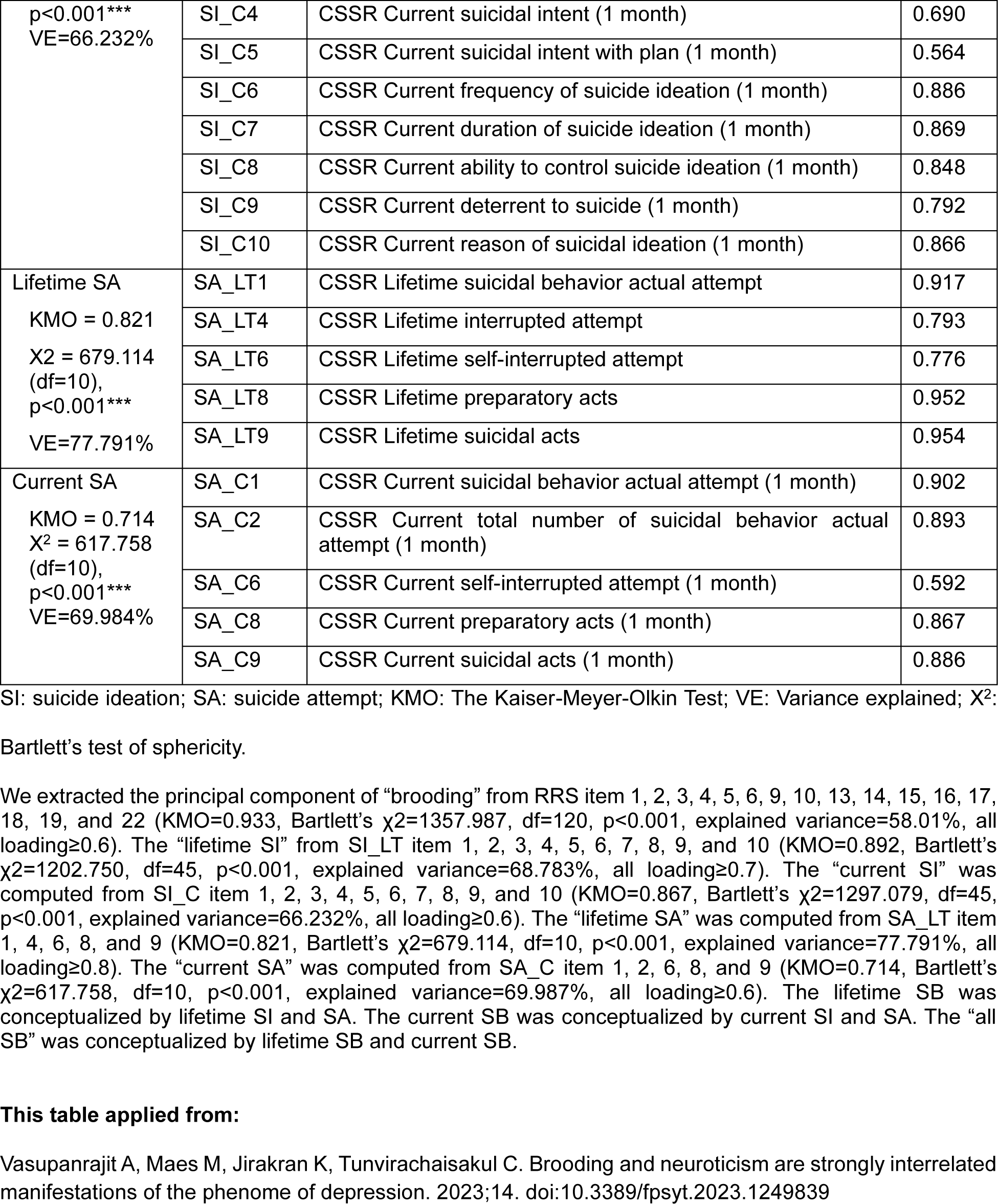
Principal components (PCs) analysis of suicide based on the items of The Columbia–Suicide Severity Rating Scale (C-SSRS) and brooding based on the items of the Ruminative Response Scale (RRS) rating scale.

